# Socio-economic disparities and predictors of fertility among adolescents aged 15 to 19 in Zambia: Evidence from the Zambia demographic and health survey (2018)

**DOI:** 10.1101/2023.12.05.23299479

**Authors:** Samson Shumba, Vanessa Moonga, Thomas O. Miyoba, Stephen Jere, Jessy M. Nkonde, Peter Mumba

## Abstract

**Background:** Annually, 12 million girls aged 15-19 give birth globally, with Africa hosting 19% of the youth aged 15-24. Zambia sees 29% of adolescents experiencing childbirth, with notable variations among age groups. Predictors of adolescent fertility include age, residential area, education, contraceptive use, and socioeconomic status. Studies emphasize increased health risks for adolescent mothers, including eclampsia and systemic infections, while infants face elevated risks such as low birth weight and severe neonatal conditions. Projections anticipate a continued rise in these trends by 2030.

**Method:** The analysis utilized the 2018 Zambia Demographic Health Survey (ZDHS). The association between dependent and independent variables was assessed using the Rao–Scott Chi-square test. Determinants of adolescent fertility were identified through a multilevel ordinal logistic regression conducted at a significance level of 5%. Graphs were generated using Excel, and the analysis was carried out using Stata version 14.2.

**Results:** A total of 3,112 adolescents were involved in the study, revealing that 75.88% had not given birth, 21.14% had one child, and 2.98% had at least two children. The findings revealed that education played a protective role, with adjusted odds ratios (AOR) of 0.47 (95% CI, 0.23 – 0.97), 0.21 (95% CI, 0.10 – 0.47), and 0.03 (95% CI, 0.00 – 0.54) for primary, secondary, and tertiary education, respectively. On the other hand, certain factors were associated with an elevated risk of fertility. These included the age of adolescents, educational attainment, marital status, wealth index, contraceptive use, exposure to family planning (FP) messages, being educated about FP at health facilities, and age at first sexual encounter. Among contextual factors, only community age at first birth was identified as a predictor of fertility, AOR, 1.59 (95% CI, 1.01 – 2.52).

**Conclusion:** The study highlights sociodemographic disparities in adolescent fertility, emphasizing the need for targeted sexual reproductive health policies. Education protects against having more than one child, while marital status significantly influences fertility, particularly for married adolescents. The research provides valuable insights into the complex factors shaping adolescent fertility in Zambia, offering guidance for interventions and policies to support this vulnerable demographic.

## Background

Every year, an estimated 21 million girls aged 15 to 19 years in developing regions become pregnant and approximately 12 million of them give birth [1]. Globally, adolescent birth rate has decreased from 64.5 births per 1000 women (15 – 19 years) in 2000 to 41.3 births per 1000 women in 2023. However, rates of change have been uneven in different regions of the world with the sharpest decline in Southern Asia and slower declines in the Latin American and Caribbean (LAC) and Sub-Saharan Africa (SSA) regions. Although declines have occurred in all regions, SSA and LAC continue to have the highest rates globally at 99.4 and 52.1 births per 1000 women, respectively in 2022. In the WHO African Region, the estimated adolescent birth rate was 97 per 1000 adolescent girls in the European Region [2].

Africa is home to the world’s youngest and rapidly growing population. In 2019, the continent had an estimated 230 million young people aged between 15 and 24, constituting 19% of the global youth population. Projections indicate that by 2030, the number of youths residing in Africa will have surged by 42 percent in 2023 [3]. Alarming trends suggest that the total number of teenage pregnancies is expected to increase by 2030, with sub-Saharan Africa projected to witness a higher prevalence. Notably, the African nations with the highest prevalence of teenage pregnancies include Niger, Mali, Angola, Mozambique, Guinea, Chad, and Cote d’Ivoire [4]. This is particularly concerning given that the region already leads in both teenage pregnancies and child marriages [5].

For more than four decades, Zambia, has grappled with elevated fertility rates [6, 7]. Over time, there has been a notable decline in the total fertility rate, dropping from 6.5 children in 1992 to 4.7 children in 2018 [7, 8]. Approximately 29% of adolescents aged 15 to 19 have already experienced childbirth, with 6% of these births occurring among 15-year-olds and a significant 58% among those aged 19 [6]. Furthermore, there has been notable variations in the percentage of adolescent girls aged 15 to 19 who have begun child bearing ranged from 14.9% in Lusaka to 42.5% in the Southern Province in 2018 [7].

Various studies have shown that the key determinants of fertility among the older adolescents and younger adults is individual’s current age, type of residential area, educational attainment, contraceptive utilization, and socioeconomic status. Child marriages deprive adolescent girls of their sexual and reproductive health rights and curtails opportunities for them to realize their full potential and enjoyment of human rights entitlements as enshrined in various international treaties [9]. In Zambia, the prevalence of child marriages stands among the highest in the world, though it has marginally decreased from 31.7% in 2014 to 29% in 2018 [10]. Disturbingly, statistics reveal that 16.5% of girls aged 15 to 19 are married, while 31.4% of those aged 20 to 24 tied the knot before reaching the age of 18 [11].

Studies have shown that adolescent mothers (aged between 10 and 19) face higher risks of eclampsia, puerperal endometritis and systemic infections than women aged 20 to 24 years, and babies of adolescent mothers face higher risks of low birth weight, preterm birth and severe neonatal condition. Moreover, research has underscored the correlation between age and maternal mortality, with a prevalence rate of 13% observed among individuals aged between 10 and 19 years [12]. Preventing pregnancy among adolescents and pregnancy related mortality and morbidity are foundational to achieving positive health outcomes across the life course and imperative for achieving the sustainable development goals (SDGs) related to maternal and newborn health [13]. In light of these concerning trends, this study aims to investigate the socio-economic disparities and associated factors of fertility among adolescents aged 15 to 19 years in Zambia.

## Methods

This study constitutes a secondary analysis of microdata utilizing national-level data sourced from the Zambia Demographic and Health Survey (ZDHS) program. The ZDHS is a comprehensive, nationally representative household survey conducted by the Zambia Statistics Agency in collaboration with global partners, including ICF International and the United States Agency for International Development (USAID). The survey employs a two-stage sampling process, initially selecting enumeration areas (EAs) and subsequently households. The nature of the DHS data enables the comparison of variables over time, facilitating the monitoring of changes in indicators across various geographical regions [14].

Participation in the survey was limited to women aged 15–19 years from selected households who had consented to take part in the research. Detailed methods employed in the DHS are comprehensively documented elsewhere [7]. For this specific study, we extracted all pertinent variables from the women’s data files (individual recode) 2018 ZDHS datasets. The data under examination pertains to the population of adolescent’s aged 15–19 years. Data collection took place from 18 July 2018 to 24 January 2019 [14]. Data was accessed 1^st^ of October and 20^th^ of November 2023. The authors in this study did not have access to information that could identify individual participants during or after data collection.

### Dependent and independent variables

The variable of interest in this study is “ever given birth/children ever born” among adolescents aged 15 to 19 years. The explanatory variables that were used in the study were demographic, socio-economic, behavioral and community level factors. The study utilized the Bongaarts proximate determinants of fertility to adopt the explanatory variables as they relate to adolescents aged 15 to 19 years [15].

Community-level variables in this study were derived by aggregating individual-level data into clusters and encompassed community poverty, community education, community knowledge of family planning (FP) methods, and place of residence. These community-level variables were dichotomized as either ‘low’ or ‘high,’ reflecting the extent of the phenomena under investigation at the cluster level. Place of residence and geographical region were maintained in their original categorizations. Place of residence played a pivotal role in the sample design, as it was utilized as a criterion to estimate the prevalence of key demographic and health indicators at the national level. It was categorized as either ‘rural’ or ‘urban’ and directly contributed to the description of community characteristics.

### Data Analysis

For descriptive purposes, frequencies and percentages were computed for categorical variables. To determine association between the outcome variable (“ever given birth”) and the categorical variables, the Uncorrelated Design Based Chi-square test (Rao – Scott Chi-square test) was used. The study furthermore utilized the survey multilevel mixed effect ordinal logistic regression (proportional odds model) to determine the factors associated with fertility among adolescents. This approach accounted for the hierarchical structure of the data, with women nested within households and households nested within clusters. The outcome variable (ever given birth; no child, 1 child and 2+ children) is ordinal and using multinomial regression would give less efficient estimates. The study used an investigator led approach, all variables were selected from wide range literature. The probability F-test showed that the adopted model explained the outcome better than the null model (model without the explanatory variables), P<0.0001. In addition, the Brant test was utilized to examine whether the model had fulfilled the parallel lines assumption. A significant test statistic would suggest a violation of the parallel regression assumption. Consequently, the proportional odds model (p=0.6083) was adopted with confidence, as it met the assumption, as illustrated in the table 3 below. The log likelihood ratio test, Akaike Information Criteria (AIC) and the Bayesian Information Criteria (BIC) were sufficiently explored to select the best fit model (see table 3). The variance for the predictors was less than 5 imposing no worry on multicolinearity in the model. Stata version 14.2 was used for the analysis.

The proportional odds model is expressed in the logit form as:

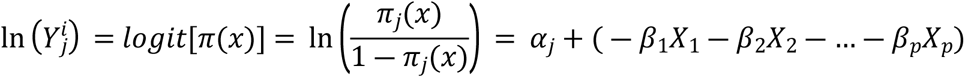

Where, *π*_*j*_(*x*) = *π*(*Y* ≤ *j*|*x*_1,_*x*_2,_…,*x*_*p*_), which is the probability of being at or below category j given a set of predictors, *j* = 1, 2,…, *j* – 1. *⍺* are the cut of points and *β*_1_,*β*_2_,…,*β*_*p*_ are logit coefficients. To estimate the ln(odds) of being at or below the *jth* category, the Proportional Odds model can be rewritten as:

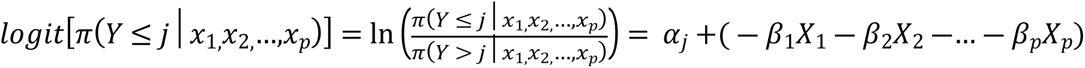

This model is based on the assumption of consistent effects represented by β for each logit [16, 17]. To find the magnitude and presence of relationships, odds ratios and their corresponding 95% confidence intervals were computed. The proportionality of odds for the dependent variable was evaluated using the Brant Test.

**Table S1:** Multicolinearity test.

| Variables | Variance Inflation Factor |
| --- | --- |
| Age of Adolescent | 1.54 |
| Education Level | 1.64 |
| Employment Status | 1.31 |
| Marital Status | 1.40 |
| Wealth Index | 3.25 |
| Contraceptive Use | 1.24 |
| Exposure to FP messages | 1.25 |
| Taught FP at H/F | 1.17 |
| Age at first sex | 1.54 |
| Frequency of listening to Radio | 1.14 |
| Frequency of listening to television | 1.83 |
| Residence | 1.89 |
| Community Poverty | 2.53 |
| Community Education | 1.85 |
| Community age at first birth | 1.03 |
| Community access to FP messages | 1.32 |
| Community employment status | 1.33 |
| Community ideal number of children | 1.10 |

### Model Selection

**Model 1** (competing model): Null model that is model without explanatory variable

**Model 2** (Competing model): Individual level factors: Age of respondent, education, employment, contraceptive, exposure to FP, marital status, wealth index, taught FP at frequency of listening to a radio and frequency of listening to a television.

**Model 3** (accepted Model): Individual level factors: Age of respondent, education, employment, contraceptive, exposure to FP, marital status, wealth index, taught FP at frequency of listening to a radio and frequency of listening to a television, residential area, community poverty, community level of education, community age at first birth, community access to FP message, community employment and community ideal number of children.

### Ethical Consideration

The DHS data survey protocols undergo a rigorous review process to ensure compliance with ethical standards. Ethical clearance for the study was granted by the ICF Institutional Review Board (IRB) under the ICF Project Number: 132989.0.000.ZM.DHS.02. The ICF IRB focuses on ensuring adherence to regulations safeguarding human subjects, while the host country IRB ensures alignment with national laws and norms.

Furthermore, the consent process includes obtaining informed oral consent from each respondent, and for adolescents under 18 years, consent is obtained from a parent or guardian. Comprehensive information about the DHS consent process is available at https://www.dhsprogram.com/What-We-Do/Protecting-the-Privacy-of-DHS-Survey-Respondents.cfm. It is crucial to highlight that authorization to use the ZDHS data was obtained from ICF Macro, and the dataset, titled ZMIR71DTA, can be accessed at https://www.dhsprogram.com/data. The user diligently followed the provided instructions, emphasizing the confidential nature of the data and the importance of not attempting to identify any household or individual respondent interviewed in the survey (ensuring anonymity).

## Results

The study’s findings reveal several notable patterns among the participants. Firstly, it is evident that a higher proportion of participants (95.61%) with no prior history of children were 15 years old. In contrast, a majority (37.26%) of participants who had two or more children were 19 years old. Furthermore, the data demonstrates that the majority of adolescents who gave birth between the ages of 15 to 17 comprised those who had only one or more children. Additionally, a significant proportion of these adolescents with one or at least children originated from rural areas (25.62% and 4.34% respectively). In terms of marital status, 60.2% of adolescents with a history of one childbirth were married, as opposed to those who were not. Similarly, a majority of adolescents with a history of multiple childbirths were married, accounting for 14.86%%. Moreover, the study’s findings suggest a significant prevalence of adolescents from low-income households (poorest) and an elevated risk of having multiple children (6.12%), in contrast to those from wealthier (richest) backgrounds (0.28%). This association was found to be statistically significant, with a p-value of less than 0.0001 as shown in table 2 below.

**Table 2:**
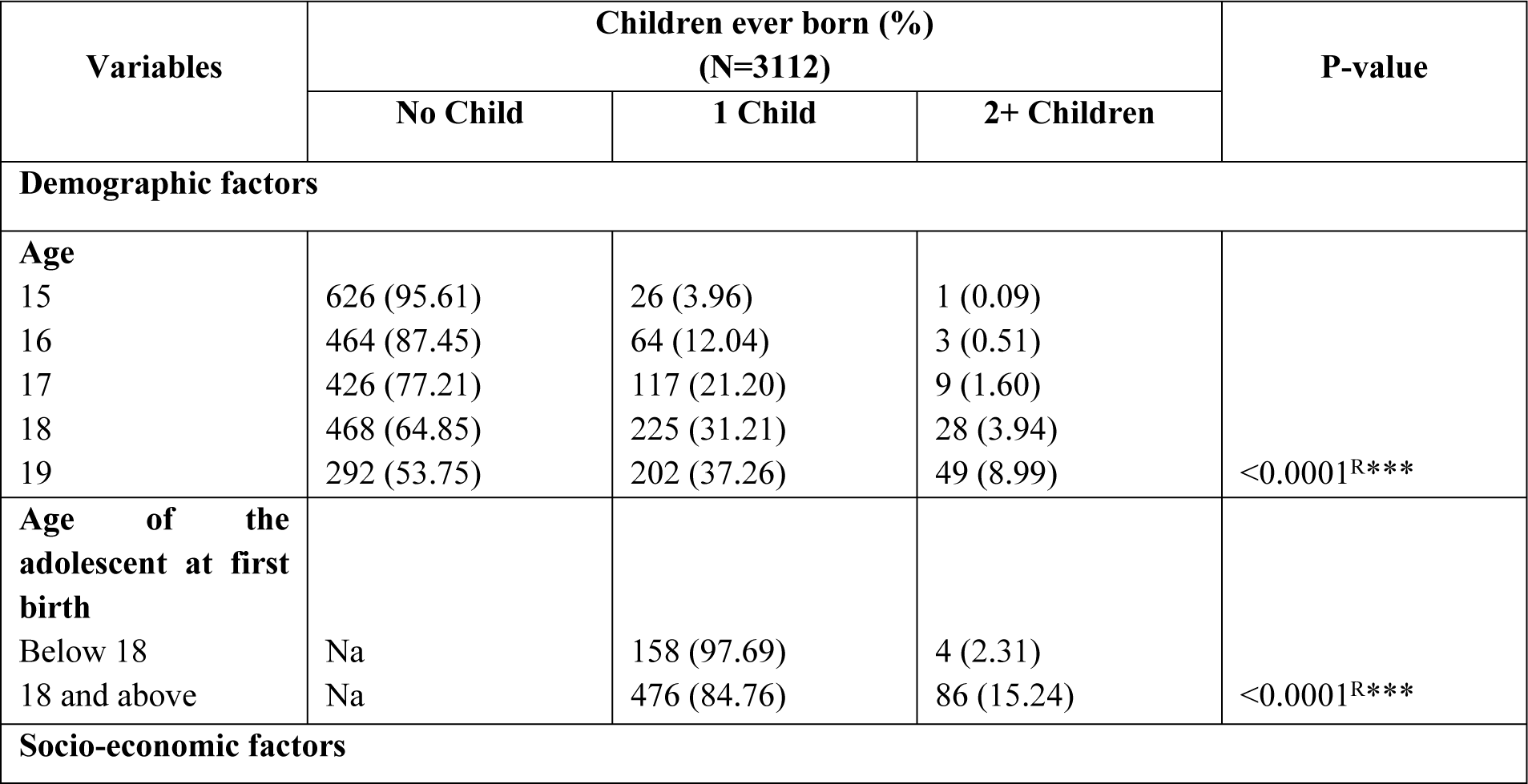
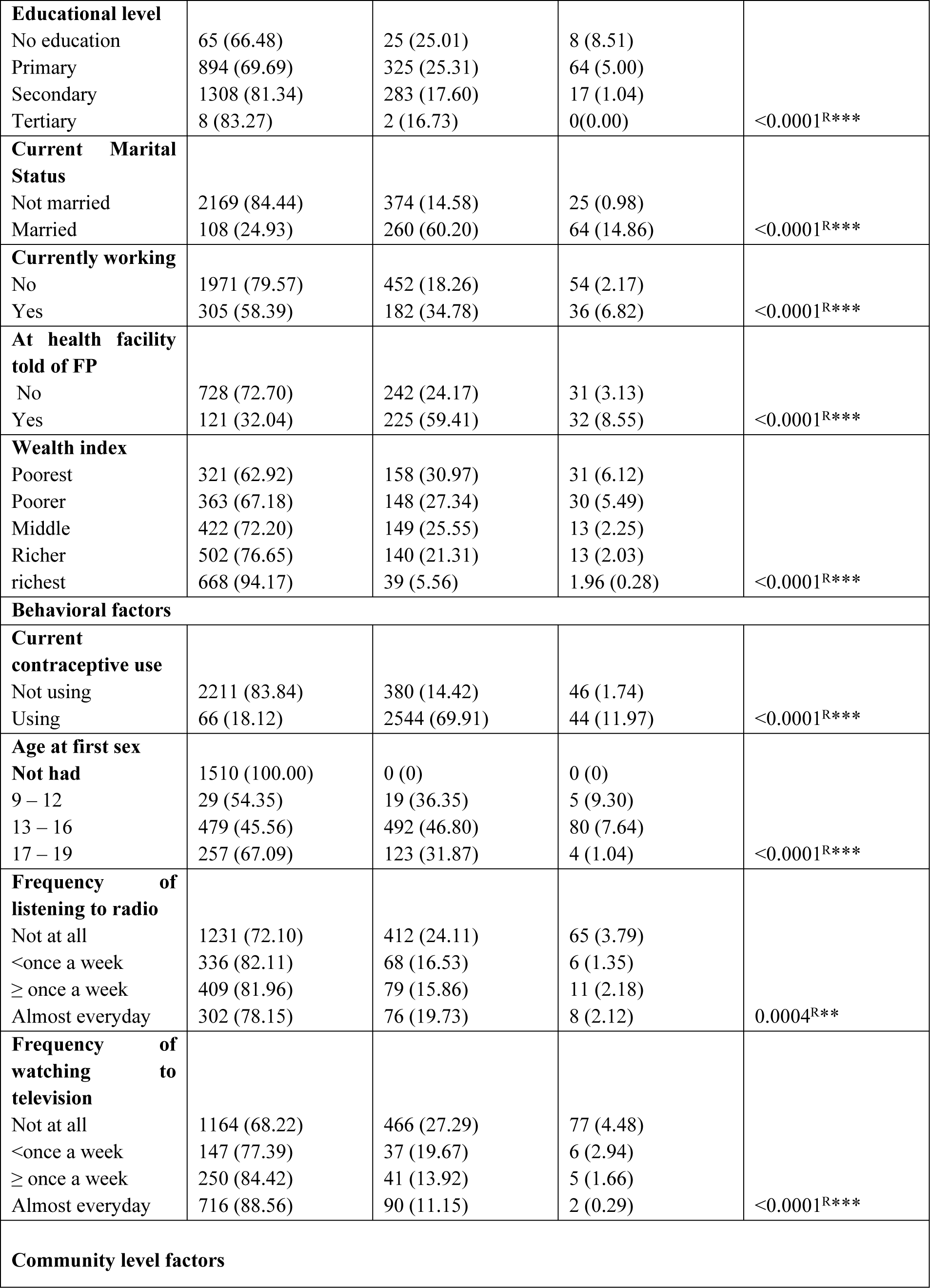
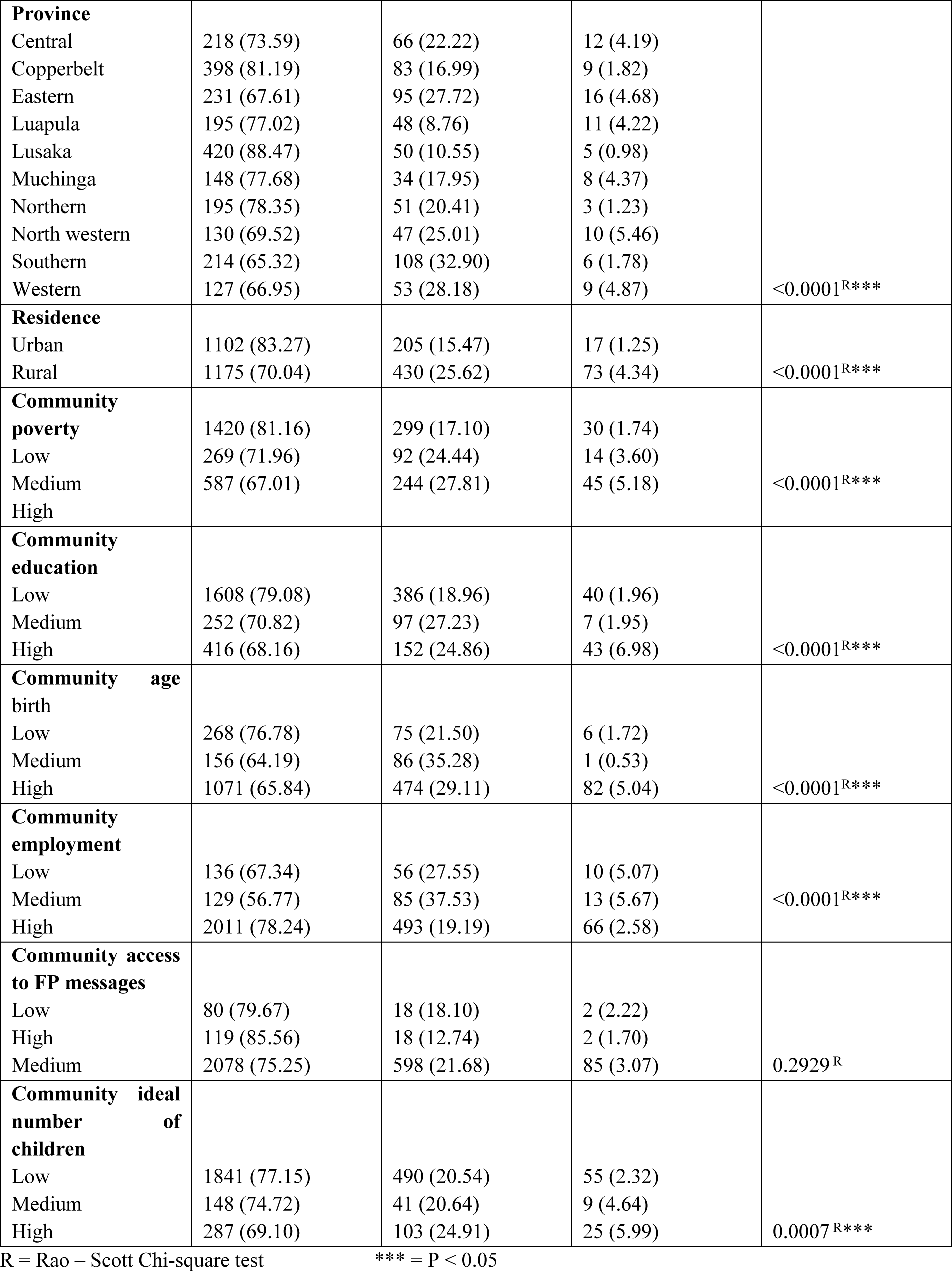
Variables distribution and association of adolescent fertility in Zambia 2018.

### Adolescent girls with no history of a childbirth

In urban settings, the prevalence of adolescents without a history of childbirth was slightly higher than in rural settings, with the exception of the southern province. In the southern province, a notable proportion of adolescent girls in rural areas reported no history of childbirth compared to their urban counterparts. Additionally, a significant difference in the population of adolescents without a history of childbirth was observed in the eastern province, as depicted in Fig1 below.

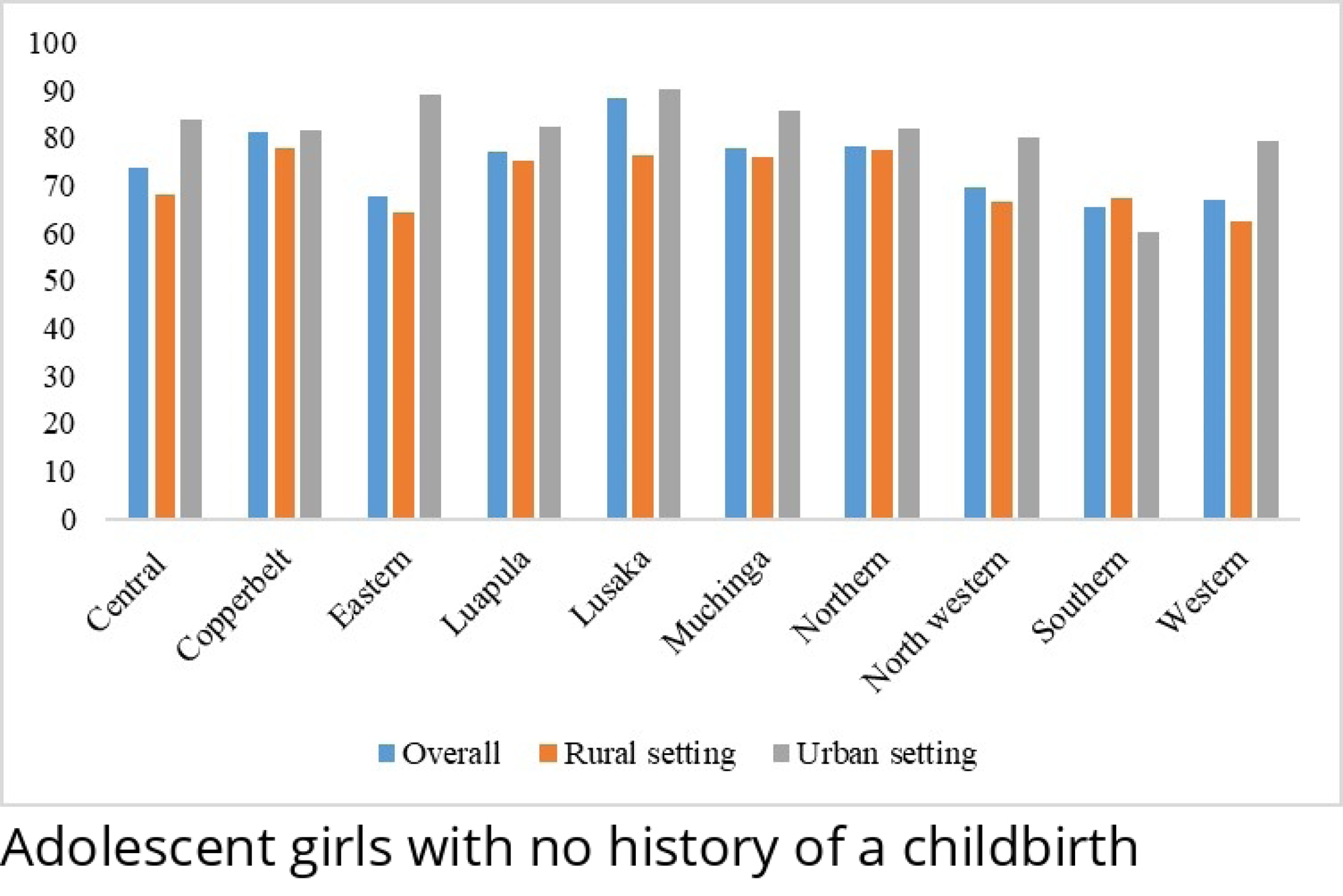

### Adolescent girls with a history of one childbirths

Additionally, the study’s findings indicate a higher count of adolescents with a history of one childbirth hailing from rural settings, with the exception of the southern province, where there is a greater proportion of adolescents in urban settings who have experienced childbirth. Furthermore, Luapula province documented the lowest percentage of adolescents with a history of one childbirth, and in urban areas, the lowest proportion was observed in Lusaka province as shown in Fig2 below.

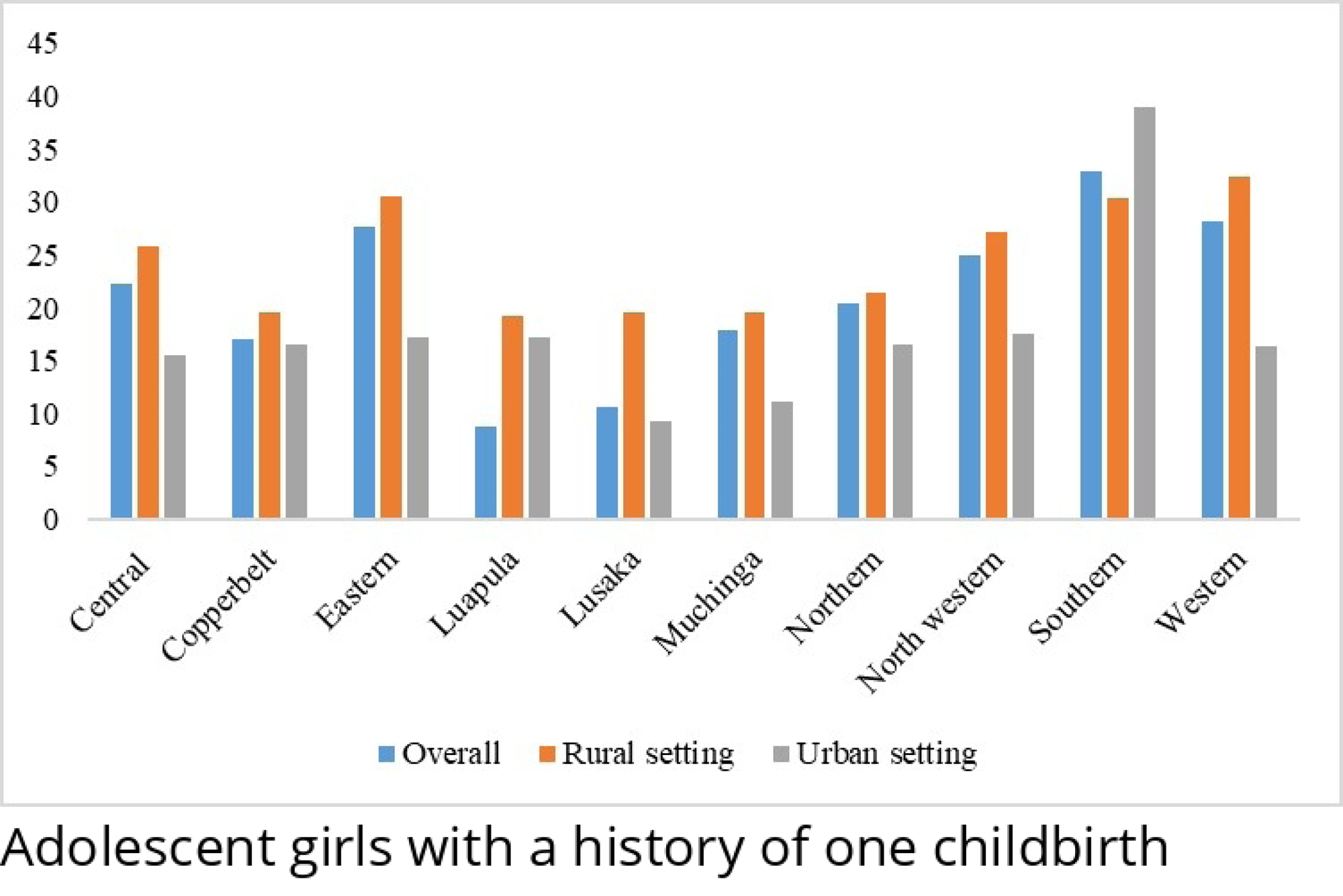

### Adolescent girls with history of at least two childbirths

The study’s results in Fig3 below reveal a higher percentage of adolescents in rural settings with a history of at least two childbirths across all provinces, except for the Northern Province. Noteworthy variations were observed in the central, eastern, Luapula, Lusaka, and northwestern provinces. Specifically, the Northern Province exhibited a higher proportion of adolescents in rural settings with a record of at least two childbirths, followed by the central province. In urban settings, the Western Province took the lead, followed by Muchinga.

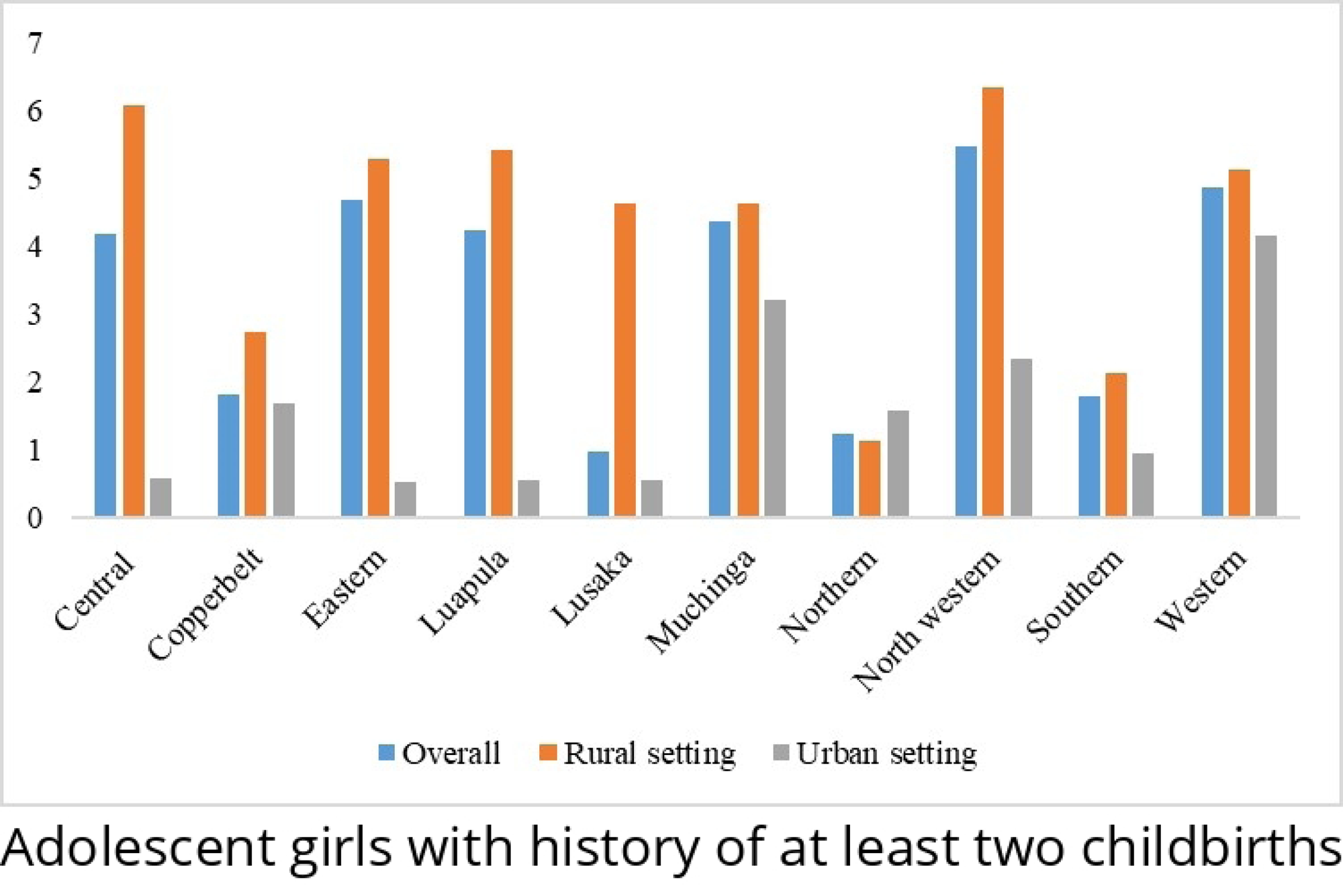

### Multilevel mixed effect ordinal logistic regression

The study investigated the factors associated with adolescent fertility using the multilevel mixed effect ordinal logistic regression. The findings suggest that a year increase in the age of adolescents increased the odds of being beyond a particular category of fertility (Multiple child births), given the effects of all other predictors are held constant. In other words, older adolescents were associated with increased odds of having multiple children, (AOR, 1.50; 95% CI, 1.31-1.72; P<0.0001) and this was statistically significant. In the same vein, adolescents with a higher level of education (primary, secondary and tertiary education) had reduced odds of being beyond a particular fertility category compared to those with no level of education (AOR, 0.47; 95% CI, 0.23 – 0.97; AOR, 0.2; 95% CI, 0.10 – 0.47; AOR, 0.03; 95% CI, 0.00 – 0.54 respectively).

Furthermore adolescents who were married, used contraceptives, coming from the poorer backgrounds or had an exposure to family planning (FP) messages had increased odds of having multiple children (being beyond a particular fertility category) (AOR, 2.56; 95% CI, 1.78 – 3.67; AOR, 3.09; 95% CI, 2.20 – 4.32; AOR, 1.68; 95% CI, 1.11 – 2.53 and AOR, 1.18; 95% CI, 1.13 – 1.22 respectively). Similarly adolescent told about FP at a health facility had increased odds of 2.77 of being beyond a particular category of fertility (higher number of births) compared to those were not told (95% CI, 2.04 – 3.76). Holding all things constant, a year increase in age at first sex increased the odds of having multiple children by a factor of 1.18 times (AOR, 1.18; 95% CI, 1.13 – 1.22). Among the contextual factors, that is residential area, community poverty, community level of education, community age at first birth, community access to FP message, community employment and community idea number of children), only community age at first birth was predicting multiple births among adolescents. In other words, adolescents from a high community age at first birth had increased odds of being beyond a particular category of fertility compared to those from a low community age at first birth (AOR, 1.59; 95% CI, 1.01 – 2.52), see table 3. Ref (1)

**Table 3:** Multilevel mixed effect ordinal logistic regression.

| Variables | Model 1<br>AOR | Model 2<br>AOR | Model 3<br>AOR |
| --- | --- | --- | --- |
| **Intercept** |  |  |  |
| Intercept 1 | 3.42 (3.07 – 3.81)*** | 8.13 (5.81 – 10.44)*** | 8.77 (6.07 – 11.47)*** |
| Intercept 2 | 3.58 (3.36 – 3.81)*** | 12.03 (9.60 – 14.46)*** | 12.78 (10.01 – 15.56)*** |
| **Socioeconomic and demographic Factors** |  |  |  |
| **Age of Adolescent** |  | 1.39 (1.22 – 1.60)*** | 1.50 (1.31 – 1.72)*** |
| **Education Level** |  |  |  |
| No Education |  | Ref (1) | Ref (1) |
| Primary |  | 0.50 (0.24 – 1.03) | 0.47 (0.23 – 0.97)* |
| Secondary |  | 0.26 (0.12 – 0.55)*** | 0.21 (0.10 – 0.47)*** |
| Tertiary |  | 0.06 (0.06 – 0.90)* | 0.03 (0.00 – 0.54)** |
| **Employment Status** |  |  |  |
| Not employed |  | Ref (1) | Ref (1) |
| Employed |  | 1.15 (0.83 – 1.60) | 1.08 (0.75 – 1.55) |
| **Marital Status** |  |  |  |
| Not married |  | Ref (1) | Ref (1) |
| Married |  | 2.49 (1.76 – 3.52)*** | 2.56 (1.78 – 3.67)*** |
| **Wealth Index** |  |  |  |
| Poorest |  | Ref (1) | Ref (1) |
| Poorer |  | 1.69 (1.13 – 2.53)* | 1.68 (1.11 – 2.53)** |
| Middle |  | 0.99 (0.65 – 1.52) | 0.86 (0.51 – 1.44) |
| Richer |  | 0.99 (0.58 – 1.70) | 0.85 (0.44 – 1.67) |
| Richest |  | 0.68 (0.33 – 1.42) | 0.74 (0.30 – 1.81) |
| **Behavioral factors** |  |  |  |
| **Contraceptive Use** |  |  |  |
| No |  | Ref (1) | Ref (1) |
| Yes |  | 3.99 (2.84 – 5.62)*** | 3.09 (2.20 – 4.32)*** |
| **Exposure to FP messages** |  |  |  |
| No |  | Ref (1) | Ref (1) |
| Yes |  | 0.62 (0.17 – 2.29) | 1.18 (1.13 – 1.22)*** |
| **Taught FP at H/F** |  |  |  |
| No |  | Ref (1) | Ref (1) |
| Yes |  | 2.86 (2.09 – 3.92)*** | 2.77 (2.04 – 3.76)*** |
| **Age at first sex** |  | 1.18 (1.13 – 1.22)*** | 1.18 (1.13 – 1.22)*** |
| **Frequency of listening to Radio** |  |  |  |
| Not at all |  | Ref (1) | Ref (1) |
| Less than once a week |  | 0.62 (0.39 – 0.98)* | 0.64 (0.40 – 1.02) |
| At least once a week |  | 0.87 (0.56 – 1.36) | 0.94 (0.60 – 1.49) |
| Almost everyday |  | 0.99 (0.63 – 1.54) | 0.91 (0.57 – 1.44) |
| **Frequency of listening to television** |  |  |  |
| Not at all |  | Ref (1) | Ref (1) |
| Less than once a week |  | 0.73 (0.38 – 1.40) | 0.74 (0.38 – 1.43) |
| At least once a week |  | 0.46 (0.24 – 0.87) | 0.56 (0.29 – 1.08) |
| Almost everyday |  | 0.71 (0.40 – 1.26) | 0.85 (0.47 – 1.55) |
| **Contextual Factors** |  |  |  |
| **Residential area** |  |  |  |
| Urban |  |  | Ref (1) |
| Rural |  |  | 1.02 (0.77 – 1.87) |
| **Community Poverty** |  |  |  |
| Low |  |  | Ref (1) |
| Medium |  |  | 0.89 (0.52 – 1.50) |
| High |  |  | 0.94 (0.55 – 1.60) |
| **Community Education** |  |  |  |
| Low |  |  | Ref (1) |
| Medium |  |  | 0.67 (0.41 – 1.07) |
| High |  |  | 0.73 (0.47 – 1.15) |
| **Community age at first birth** |  |  |  |
| Low |  |  | Ref (1) |
| Medium |  |  | 1.24 (0.67 – 2.28) |
| High |  |  | 1.59 (1.01 – 2.52)* |
| **Community access to FP messages** |  |  |  |
| Low |  |  | Ref (1) |
| High |  |  | 0.43 (0.11 – 1.66) |
| Medium |  |  | 0.55 (0.23 – 1.33) |
| **Community employment status** |  |  |  |
| Low |  |  | Ref (1) |
| Medium |  |  | 1.91 (0.99 – 3.97) |
| High |  |  | 1.22 (0.71 – 2.09) |
| **Community ideal number of children** |  |  |  |
| Low |  |  | Ref (1) |
| Medium |  |  | 0.77 (0.46 – 1.28) |
| High |  |  | 0.94 (0.63 – 1.39) |
| **Random effects** |  |  |  |
| Variance (SE) | 0.10 | 0.19 | - |
| VPC (%) | 0.40 | 0.31 | 0.00 |
| AIC | 3985.62 | 1534.32 | 1379.77 |
| BIC | 4003.75 | 1656.18 | 1557.63 |
| Log likelihood | -1989.81 | -744.16 | - 654.88 |
| VIF | Na | <5 | <5 |

\*\*\* = P-value ≤ 0.01
\*\* = 0.01 < P-value ≤ 0.03
\* = 0.03 < P-value < 0.05

## Discussion

This study analyzed the predictors of fertility among older adolescents aged between 15 and 19 in Zambia. Utilizing a multilevel ordinal logistic regression on data from the 2018 Zambia Demographic and Health Surveys, the study aimed to gain deeper insights into the factors contributing to elevated adolescent fertility in this age group. Disparities in the prevalence of higher adolescent fertility were noted across various sociodemographic groups. Understanding the factors linked to increased adolescent fertility in Zambia holds significant implications for enhancing sexual reproductive health policies and programs. This, in turn, contributes to the ongoing efforts to reduce teenage pregnancies and the incidence of HIV and other sexually transmitted diseases (STIs).

The study identified demographic and socio-economic disparities in adolescent fertility, revealing that a majority of adolescents with one or more children were aged 19. About 60% of married adolescents had one child, and approximately 14% in rural areas had more than one child. Rural areas showed significant increases in adolescents with multiple children. Furthermore, those with no formal education, married, residing in rural areas, low-income, or unemployed were more likely to have multiple children. This aligns with studies in Kenya, Nigeria, Malawi, and Tanzania [18–21]. However, the study noted that the percentage of adolescents with one child was higher than in Malawi, Uganda, Tanzania, Ethiopia, Rwanda, Eritrea, and Ghana [22–25].

The research revealed that certain individual factors such as education, marital status, wealth index, contraceptive use, exposure to family planning messages, and receiving family planning education at health facilities were significantly associated with higher adolescent fertility in Zambia. Notably, only community age at first birth emerged as a significant predictor of higher adolescent fertility. Among the models considered, model three, which incorporated both individual and community-level variables, was accepted. This decision was based on its superior fit, as evidenced by smaller values of AIC, BIC, and Log likelihood in comparison to the null model (without individual and community level variables) and model two (including only individual-level variables).

Furthermore, the study findings indicate that older adolescents face an increased risk of having more than one child. This aligns with observations from studies conducted in Kenya, Nigeria and Ghana, where it was noted that older adolescents are more susceptible to higher fertility compared to their younger counterparts [18, 22, 26]. A similar trend was identified in a study conducted in Indonesia, revealing a positive correlation between the age of adolescents and birthrate [27]. Similarly, our study revealed that adolescents who were older at the time of their first sexual encounter and those from communities with a higher community age at birth were more likely to have more than one child. This finding contradicts a study conducted in Uganda, which found that individuals who engaged in early sexual activity and delayed childbirth were less likely to experience higher fertility compared to those who underwent these events early [18, 28].

Our findings indicate that adolescent girls who completed primary, secondary, and tertiary education were less likely to have more than one child compared to their counterparts with no formal education. This observation is consistent with studies conducted in Nigeria, where it was established that women with primary education or higher exhibited lower risks of having children in comparison to adolescents with no formal education [20, 29]. This suggests that education plays a crucial role in enhancing autonomy, decision-making abilities, and economic independence. Consequently, it contributes to delaying marriage and sexual debut while increasing knowledge about contraception, as noted in studies such as those by Mohr et al [30].

Marital status in the study similarly predicted adolescent fertility that is the findings suggests that adolescent girls who are married had a higher chance of having multiple children compared to those who were not. This aligns with research conducted in various African countries, including Burundi, Ethiopia, Nigeria, Kenya, Congo, and Central Africa, where early marriages have been identified as a significant factor contributing to high fertility rates [18, 31, 32, 33]. This implies that early child marriages play a central role in driving increased adolescent fertility. The pressure exerted on married adolescents by their families and partners to start their own families contributes to this phenomenon. Teenage pregnancies resulting from early marriages pose heightened risks, including increased chances of maternal mortality during childbirth and elevated rates of neonatal and infant deaths among young mothers. Girls aged between 15 and 19 face double the likelihood of dying during childbirth compared to women aged 20 and above. This risk is further compounded by factors such as HIV, making complications during pregnancy and childbirth the primary cause of death for young women in this age group. Additionally, pregnant adolescents are more susceptible to serious health issues such as eclampsia, puerperal endometritis, and systemic infections compared to adults [34].

Furthermore adolescents who used contraceptives, were exposed to family planning (FP) or received FP education at health facilities were more likely to have more than one child compared to those who did not use contraceptives. This aligns with research conducted in Kenya and Zambia, which indicated that adolescents using contraceptives were at a higher risk of experiencing higher fertility [18, 35]. This phenomenon could be attributed to the observation that adolescents often initiate contraceptive use after having their first child. Furthermore, a contributing factor may be the cultural norms that discourage having children out of wedlock, particularly prevalent in rural areas, leading many girls who become pregnant to enter early marriages [36].

In contrasts to the use of contraceptives, exposure to FP or receiving FP the findings in Ethiopia, Namibia, and Burundi, where research indicated that teenagers who receive sexual education through media channels, such as television and radio, are likely to abstain from sexual intercourse due to knowledge about the dangers of early engagement, such as sexually transmitted infections and risky pregnancies leading to potential harm [31, 37, 38]. This inconsistency might be attributed to the perceptions held by young people that family planning messages are primarily intended for older married individuals and do not apply to them, leading them to disregard or avoid such messages [32].

Residence did not emerge as a significant predictor of adolescent fertility in the study, a result that aligns unexpectedly with studies conducted in Ethiopia, Uganda, and Mexico. These studies similarly found that the place of residence was not a significant predictor of higher adolescent fertility in Zambia [39, 40]. In contrasts to the findings in the descriptive results that shows a higher proportion of adolescents with at least one childbirth residing in rural areas across majority of the provinces. The differences are much wider among adolescents with at least two multiple childbirths.

### Limitations and justification of the study

This study draws strength from the utilization of national data, providing a representative sample of the adolescent female population aged 15 to 19 in Zambia. Consequently, the study’s findings are applicable and can be generalized to the specified target population of adolescent girls within this age range. However, it is essential to acknowledge the study’s limitations. The reliance on the latest Zambia Demographic and Health Survey (ZDHS) dataset from 2018 follows a cross-sectional study design, implying that the results indicate correlation rather than causation between the outcome of interest and individual or contextual factors. Additionally, caution is advised when extending the findings to the broader adolescent age group of 10 to 19 years. Moreover, the contextual factors utilized in the study are derived from the ZDHS, potentially limiting their ability to fully capture the community experience. These considerations are crucial for a nuanced interpretation of the study’s outcomes.

### Recommendation and Conclusion

The study has shown disparities in adolescent fertility across sociodemographic groups, and further emphasizing the importance of understanding contributing factors for effective sexual reproductive health policies. Education emerged as a protective factor, with completion of primary, secondary, and tertiary education associated with lower likelihoods of having more than one child. Marital status played a significant role, with married adolescent girls having a higher likelihood of multiple children, highlighting the impact of early marriages on fertility rates. Residence did not predict adolescent fertility. Furthermore, the study provided valuable insights into the multifaceted nature of adolescent fertility in Zambia, emphasizing education, marital status, and considerations of contraceptive use and cultural influences.

Furthermore the recommendations are to implement education-focused initiatives, including awareness campaigns, to promote primary, secondary, and tertiary education for adolescents. Develop strategies to prevent early marriages through legal reforms and community awareness. Enhance reproductive health education with accurate information on contraception, family planning, and consequences of early childbearing. Tailor community-based interventions to address contextual factors, engaging local leaders and considering cultural nuances. Support ongoing research and monitoring to inform policies and ensure effectiveness. A comprehensive, multifaceted approach can effectively reduce adolescent fertility and improve well-being, especially in rural areas.

## Data Availability

authorization to utilize the data was secured from ICF Macro, accessible at https://dhsprogram.com/data, under the dataset title ZMIR71DTA

## Acknowledgements

We wish to express our sincere gratitude to the Zambia Statistics Agency and the DHS Program for granting permission to utilize the 2018 ZDHS dataset.

## Notes

### Competing Interest Statement

The authors have declared no competing interest.

### Funding Statement

No funding

### Author Declarations

authorization to utilize the data was secured from ICF Macro, accessible at https://dhsprogram.com/data

## Reference

1. Sully EA, Biddlecom A, Daroch J, Riley T, Ashford L, Lince-Deroche N, Lauren Firestein and Rachel Murro. Adding It Up: Investing in Sexual and Reproductive Health 2019. Guttmacher Inst. 2020.

2. United Nations Department of Economic and Social Affairs. World Population Prospects, 2019 Revision: Age-specific fertility rates by region, subregion and country, 1950-2100 (births per 1,000 women) Estimates. Online Ed. 2021 Dec;

3. Mo Ibrahim Foundation. Ibrahim Forum Report: Africa’s youth: jobs or migration? Demography, economic prospects and mobility. 2019. https://mo.ibrahim.foundation/sites/default/files/2020-05/2019-forum-report_0.pdf.

4. WorldAtlas. Highest Teen Pregnancy Rates Worldwide. 2021; https://www.worldatlas.com/articles/highest-teen-pregnancy-rates-worldwide.html.

5. Ahinkorah BO, Kang M, Perry L, Brooks F, Hayen A. Prevalence of first adolescent pregnancy and its associated factors in sub-Saharan Africa: A multi-country analysis. Ortega JA, editor. PLOS ONE. 2021 Feb 4; 16(2):e0246308.

6. Central Statistics Office (CSO). Demographic and Health Survey Key Indicators, in Demographic and Health Survey. 2019. The DHS Program: Central Statistical Office, Lusaka.

7. Bakibinga P, Matanda D, Kisia L, Mutombo N. Factors associated with use of injectables, long-acting and permanent contraceptive methods (iLAPMs) among married women in Zambia: analysis of demographic and health surveys, 1992–2014. Reprod Health. 2019 Dec; 16(1):78.

8. Phiri M, Banda C, Lemba M. Why is Zambia’s rural fertility declining at slow pace? A review of DHS data 1992–2018. Int J Res Publ Rev. 2020 2:5–16;

9. Population Council, United Nations Population Fund (UNFPA), Government of the Republic of Zambia. Child marriage in Zambia. Lusaka, Zambia: Population Council, UNFPA and Government of the Republic of Zambia. 2017;

10. World Vision. A situation report on child marriages in Zambia. 2022; https://www.wvi.org/zambia/article/situation-report-child-marriages-zambia

11. Central Statistics Office (CSO). Demographic and Health Survey Key Indicators, in Demographic and Health Survey. 2019. The DHS Program: Central Statistical Office, Lusaka. 2014;

12. Chikonde Musonda N, Chola M, Kaonga P, Shumba S, Jacobs C. Trends and Associated Factors of Maternal Mortality in Zambia: Analysis of Routinely Collected Data (2015-April 2019). J Gynecol Obstet. 2021; 9(5):155.

13. World Health Organization. Adolescent pregnancy. 2023;

14. Croft TN, Marshall, AMJ, Allen CK. Guide to DHS Statistics. Rockv Md USA ICF. 2018; https://www.DHSprogram.com

15. Bongaarts J. A Framework for Analyzing the Proximate Determinants of Fertility. Popul Dev Rev. 1978 Mar; 4(1):105.

16. Obare F, Odwe G, Birungi H. Adolescent sexual and reproductive health situation: Insights from the 2014 Kenya Demographic and Health Survey [Internet]. Population Council; 2016 [cited 2023 Nov 21]. Available from: https://knowledgecommons.popcouncil.org/departments_sbsr-rh/251

17. Agresti A. An introduction to categorical data analysis: John Wiley & Sons. 2018;

18. Monari N, Orwa J, Agwanda A. Adolescent fertility and its determinants in Kenya: Evidence from Kenya demographic and health survey 2014. Atiqul Haq SM, editor. PLOS ONE. 2022 Jan 12; 17(1):e0262016.

19. Ngoda OA, Renju J, Mahande MJ, Kagoye SA, Mboya IB, Msuya SE. Trends and Factors Associated with Adolescent Pregnancies in Tanzania from 2004-2016: Evidence from Tanzania Demographic and Health Surveys. East Afr Health Res J. 2023 Jul 12;7(1):40–8.

20. Akombi-Inyang BJ, Woolley E, Iheanacho CO, Bayaraa K, Ghimire PR. Regional Trends and Socioeconomic Predictors of Adolescent Pregnancy in Nigeria: A Nationwide Study. Int J Environ Res Public Health. 2022 Jul 5; 19(13):8222.

21. Palamuleni ME. Determinants of adolescent fertility in Malawi. Gender and Behaviour. 2017 Dec 1;

22. Nyarko SH. Determinants of adolescent fertility in Ghana. International Journal of Sciences: Basic and Applied Research. 2012 1):21–32;

23. Loaiza E, Liang M. Adolescent pregnancy: a review of the evidence: Unfpa; 2013;

24. Davis K, Blake J. Social structure and fertility: An analytic framework. Economic development and cultural change. 1956 Apr 1;

25. Mahy M, Gupta N. Trends and differentials in adolescent reproductive behavior in Sub-Saharan Africa: ORC Macro, MEASURE DHS; 2002;

26. Determinants of Adolescent Childbearing in Oghara Kingdom, Delta State, Nigeria. J Cult Soc Dev [Internet]. 2020 Jan [cited 2023 Nov 21]; Available from: https://www.iiste.org/Journals/index.php/JCSD/article/view/51088

27. Raharjo BB, Nugroho E, Cahyati WH, Najib N, Nisa AA. Proximate determinant of adolescents’ fertility in Central Java. 2019;

28. Gideon R. Factors associated with adolescent pregnancy and fertility in Uganda: analysis of the 2011 demographic and health survey data. American Journal of Sociological Research. 2013;3(2):30–5.

29. Maharaj NR. Adolescent pregnancy in sub-Saharan Africa – a cause for concern. Front Reprod Health. 2022 Dec 2; 4:984303.

30. Mohr R, Carbajal J, Sharma BB. The Influence of Educational Attainment on Teenage Pregnancy in Low-Income Countries: A Systematic Literature Review. J Soc Work Glob Community [Internet]. 2019 Dec 17 [cited 2023 Nov 21]; 4(1). Available from: https://scholarworks.waldenu.edu/jswgc/vol4/iss1/2

31. Liga AD, Boyamo AE, Jabir YN, Tereda AB. Prevalence and correlates associated with early childbearing among teenage girls in Ethiopia: A multilevel analysis. Tenaw LA, editor. PLOS ONE. 2023 Aug 8; 18(8):e0289102.

32. Nibaruta JC, Kamana B, Chahboune M, Chebabe M, Elmadani S, Turman JE, et al. Prevalence, trend and determinants of adolescent childbearing in Burundi: a multilevel analysis of the 1987 to 2016–17 Burundi Demographic and Health Surveys data. BMC Pregnancy Childbirth. 2022 Sep 1; 22(1):673.

33. Somefun OD, Olamijuwon E. Community structure and timing of sexual activity among adolescent girls in Nigeria. Ortega JA, editor. PLOS ONE. 2022 Jul 27; 17(7):e0269168.

34. Population Council, United Nations Population Fund (UNFPA), Government of the Republic of Zambia. Child marriage in Zambia. Lusaka, Zambia. 2021;

35. Munakampe MN, Fwemba I, Zulu JM, Michelo C. Association between socioeconomic status and fertility among adolescents aged 15 to 19: an analysis of the 2013/2014 Zambia Demographic Health Survey (ZDHS). Reprod Health. 2021 Dec; 18(1):182.

36. Sánchez-Páez DA, Ortega JA. Adolescent contraceptive use and its effects on fertility. Demographic Research. 2018 Jan 1; 38:1359–88.

37. University of Namibia, Shinyemba T, Oyedele O, University of Namibia, Kazembe L, University of Namibia, et al. Assessing the Impact of Proximate and Non-Proximate Determinants of Fertility in Namibia: A Structural Equation Modelling Approach. Namib J Res Sci Technol. 2022 May 1; 21–30.

38. Mohammed S. Analysis of national and subnational prevalence of adolescent pregnancy and changes in the associated sexual behaviours and sociodemographic determinants across three decades in Ghana, 1988–2019. BMJ Open. 2023 Mar; 13(3):e068117.

39. Chandra-Mouli V, Ferguson BJ, Plesons M, Paul M, Chalasani S, Amin A, et al. The Political, Research, Programmatic, and Social Responses to Adolescent Sexual and Reproductive Health and Rights in the 25 Years since the International Conference on Population and Development. J Adolesc Health. 2019 Dec; 65(6):S16–40.

40. Kassa GM, Arowojolu AO, Odukogbe ATA, Yalew AW. Trends and determinants of teenage childbearing in Ethiopia: evidence from the 2000 to 2016 demographic and health surveys. Ital J Pediatr. 2019 Dec; 45(1):153.

